# Shifts in Clinical Practice-Changing Acute Ischemic Stroke Research Over the Last Decade

**DOI:** 10.64898/2026.03.22.26349031

**Authors:** Mahnoor Khalid, Connor Nguyen, Jack Li, Aditya Bala, Tudor G. Jovin, Ashutosh P. Jadhav, Ngoc Mai Le, Javier Gomez Farias, Freya Kanakhara, Eunyoung A. Lee, David S. Liebeskind, Joseph N. Samaha, Hussain Azeem, Bruna Kfoury, Aman Yarlagadda, Sunil A. Sheth

## Abstract

**Background:** The past decade has witnessed rapid growth of clinical-trial programs in Europe and Asia, with randomized clinical trials (RCTs) publications from these regions outpacing those of the U.S. However, limited data exist quantifying their relative influence on practice-defining results. Here, we evaluate these shifts by analyzing geographic origin, funding source, and clinical impact of practice-changing RCTs.

**Methods:** From the 2018 and 2026 American Heart Association/American Stroke Association (AHA/ASA) Acute Ischemic Stroke (AIS) Guidelines, we identified RCTs supporting new recommendations and extracted geographic origin (China/Europe/USA/Other), funding source (government/academic/non-profit vs. industry (private/mixed); NIH vs. non-NIH), and research topic (endovascular therapy (EVT), thrombolysis, imaging, poststroke care, and prehospital and systems of care). Analyses used unweighted, reference-density-weighted, and clinical-impact-weighted strategies. Temporal trends were assessed using the chi-square/Fisher’s exact tests, with Rao-Scott adjusted chi-square tests accounting for weighting.

**Results:** We identified 21 new recommendations (47 RCTs) in 2018 and 45 (89 RCTs) in 2026. In 2018, Europe led (51.1%), followed by the U.S. (31.9%), while China and other regions contributed minimally. By 2026, Europe remained first (36%), China rose to second (29.2%), and the U.S. declined to the smallest share (14.6%), across all weighted analyses (p<0.01). NIH-funded trials declined significantly from 21.3% (unweighted), 27.4% (reference-density-weighted), and 27.3% (clinical-impact-weighted) in 2018 to 4.5%, 4.8%, and 3.4%, respectively in 2026 (p<0.01 across all weighted strategies).

**Conclusion:** In this analysis, we identify a shift away from U.S.-based clinical trials and increasing contributions from China. U.S.-based RCTs fell from the second most cited to the least cited sources of practice-changing recommendations. NIH-funded research fell from nearly one-quarter in 2018 to <5% in 2026, highlighting increasing dependence on non-U.S. studies for U.S.-based care. These findings raise questions about the effectiveness of current AIS research paradigms in the U.S.

## INTRODUCTION

The global clinical research landscape has changed markedly over the past decade. Trial outputs from Europe and Asia, particularly China, have grown rapidly, with China surpassing the U.S. in both research expenditure and volume of scientific publications.^1^ These shifts have resulted from increased international research collaboration, investments in trial infrastructure, and policy efforts to strengthen research integrity.^2–4^ As a result, international trials are now being initiated and published faster than those based in the U.S.^5^

Although the U.S. remains a leading contributor in biomedical research overall, its current role as well as how the role has changed over time, remains incompletely characterized. Further, because the raw number of trials being published is not necessarily an accurate indicator of the realized clinical impact, more nuanced approaches are necessary. For example, a trial evaluating a clinical question with study design and results comparable to 4 or 5 preceding trials may add only incremental value. As such, additional metrics are needed to weigh the overall influence of these studies when ascertaining their value.

To study the global landscape of clinical-practice defining acute ischemic stroke (AIS) research, here we analyze data from a widely accepted standard-defining guideline, the American Heart Association/American Stroke Association (AHA/ASA) AIS Guidelines.^6^ Although previous studies have analyzed global trends in clinical trial registration and geographical distribution of stroke trials within the U.S., no analysis has addressed the issue of overall clinical-practice impact, particularly using multiple complementary weighting strategies.^7–9^ By analyzing the geographic origin, funding source, and clinical impact of randomized clinical trials (RCTs) that generated new recommendations in these guidelines, we identify evolving international trends in sources of evidence.

## METHODS

### Study Design and Guideline Selection

We conducted a retrospective comparative bibliometric analysis of the 2018 and 2026 editions of the AHA/ASA Guidelines for the Early Management of Patients with AIS. We evaluated temporal shifts in RCT-derived evidence supporting the new recommendations in each guideline edition. To ensure a balanced comparison between the two guidelines and to capture significant shifts in the global research landscape, the focused 2019 update to the 2018 guideline was excluded from analysis. Only the complete 2018 and 2026 guideline documents were included.

### Reference Identification and Eligibility

We identified new recommendations from the 2018 and 2026 AHA/ASA AIS guidelines that were labeled as new or revised by the writing committees. Recommendations that were previously present but only reworded for clarity or carried over without substantive changes in clinical practice were not included. For each new recommendation, all cited references were reviewed in full, and eligibility was limited to RCTs. This restriction was applied because RCTs provide the highest level of clinical evidence and greatly impact the generation, strength, and wording of guideline recommendations. Observational studies, cohort studies, registry analyses, case-control studies, meta-analyses, systematic reviews, expert opinion statements, and other non-randomized designs were excluded. References not directly supporting a newly designated recommendation were also excluded.

### Data extraction and classification

For each included RCT, geographic origin was categorized as China, Europe, United States, or Other (Countries outside the Europe, China and U.S.: Australia, New Zealand, Japan, South Korea, Canada, Chile). For multinational trials, geographic classification was based on the location of the coordinating center or the first author’s primary institutional affiliation. Funding source was determined from the acknowledgment and disclosure sections of the original publication, categorizing as government/academic/non-profit or industry (private/mixed) funding. Funding source was further stratified into National Institutes of Health (NIH) versus non-NIH sources. Each RCT was also classified into a primary research area, defined as endovascular therapy (EVT), thrombolysis, imaging, prehospital and systems of care, or poststroke care. These categories were determined by the corresponding category definitions within the guideline publication. Each eligible RCT was also categorized by the Class of Recommendation (I, IIA, IIB, III) assigned to the recommendation within which it was cited.

### Weighting Strategies

To differentiate between the raw number of RCT references and their relative influence on guideline development, we applied three analytic weighting strategies. First, we performed an unweighted analysis where each eligible RCT publication contributed equally, regardless of the total number of references cited in the recommendation it supported. Next, we conducted a reference-density-weighted analysis, in which each trial was weighted as 1 divided by the total number of RCTs cited for that recommendation to account for citation density. As an example, in this analysis, if 4 trials were cited for a single recommendation, each trial was assigned a weight of 0.25. Finally, we performed a clinical-impact-weighted analysis, in which each new recommendation was assigned a clinical impact score from 1 (minimal expected impact on clinical practice) to 5 (major expected impact on practice). To generate a clinical-impact-weighted measure for each RCT, the recommendation’s clinical impact score was multiplied by the RCT’s reference-density-weight. The clinical impact score is a subjective evaluation made by a board-certified vascular neurologist with over 15 years of experience in stroke care and involvement in guideline development. Greater scores were assigned to recommendations with higher class of recommendation, level of evidence, and their perceived influence on practice-changing recommendations. Recommendations that lead to paradigm shifts in acute therapy (i.e. expanded thrombolysis window, expanded indications for EVT), which may result in system-wide changes in care, received higher scores relative to recommendations advised against interventions that were already not routinely performed prior to publication (e.g., routine use of hypothermia in stroke or routine use of sonothrombolysis as an adjuvant to thrombolysis). The full list of recommendations, their cited RCTs, and their assigned clinical impact scores can be found in Supplementary Table 1.

### Statistical Analysis

We evaluated temporal differences between the 2018 and 2026 guidelines across geographic distribution, funding source composition, NIH versus non-NIH funding, and research area focus. All comparisons were conducted separately for unweighted, reference-density-weighted, and clinical-impact-weighted analyses. For unweighted analyses, categorical variables were compared using Pearson’s chi-square test or Fisher’s exact test as appropriate. For weighted analyses (reference-density-weighted and clinical-impact-weighted), the Rao-Scott adjusted chi-square test (second-order correction) was applied to account for non-integer cell values and the weighting structure, ensuring that p-values appropriately reflect the design effect introduced by weighting. All statistical analyses were performed using Stata (version 19.2, StataCorp LLC, College Station, TX), with a two-sided p-value of less than 0.05 considered statistically significant.

## RESULTS

We identified 21 new recommendations in the 2018 guidelines and 45 in the 2026 update. These recommendations were supported by 47 RCTs in 2018 (Class I: 6; Class IIA: 10; Class IIB: 22; Class III: 9) and 89 RCTs in 2026 (Class I: 15; Class IIA: 24; Class IIB: 24; Class III: 26) (Supplementary Table 2). The geographic distribution of supporting RCTs shifted significantly between the two guideline eras across all weighting schemes (p<0.01; Figure 1). In 2018, Europe was the largest overall contributor to the RCT evidence base (51.1%), followed by the United States (31.9%), with China and other regions each contributing 8.5%. By 2026, Europe remained the leading contributor (36%); however, its proportional share decreased relative to 2018 (-15.1%) with similar directional decreases across weighted analyses. In contrast, China showed the largest positive shifts in contribution, rising from 8.5% in 2018 to 29.2% in 2026 (+20.7%), becoming the second-largest contributor overall. Other regions also showed an increase in contribution from 8.5% to 20.2% (+11.7%). The United States was the only group with a decline in both absolute number of RCTs, as well as in overall proportional contribution (31.9% to 14.6%; −17.3%), becoming the lowest contributor across all weighting schemes.

**Figure 1.**
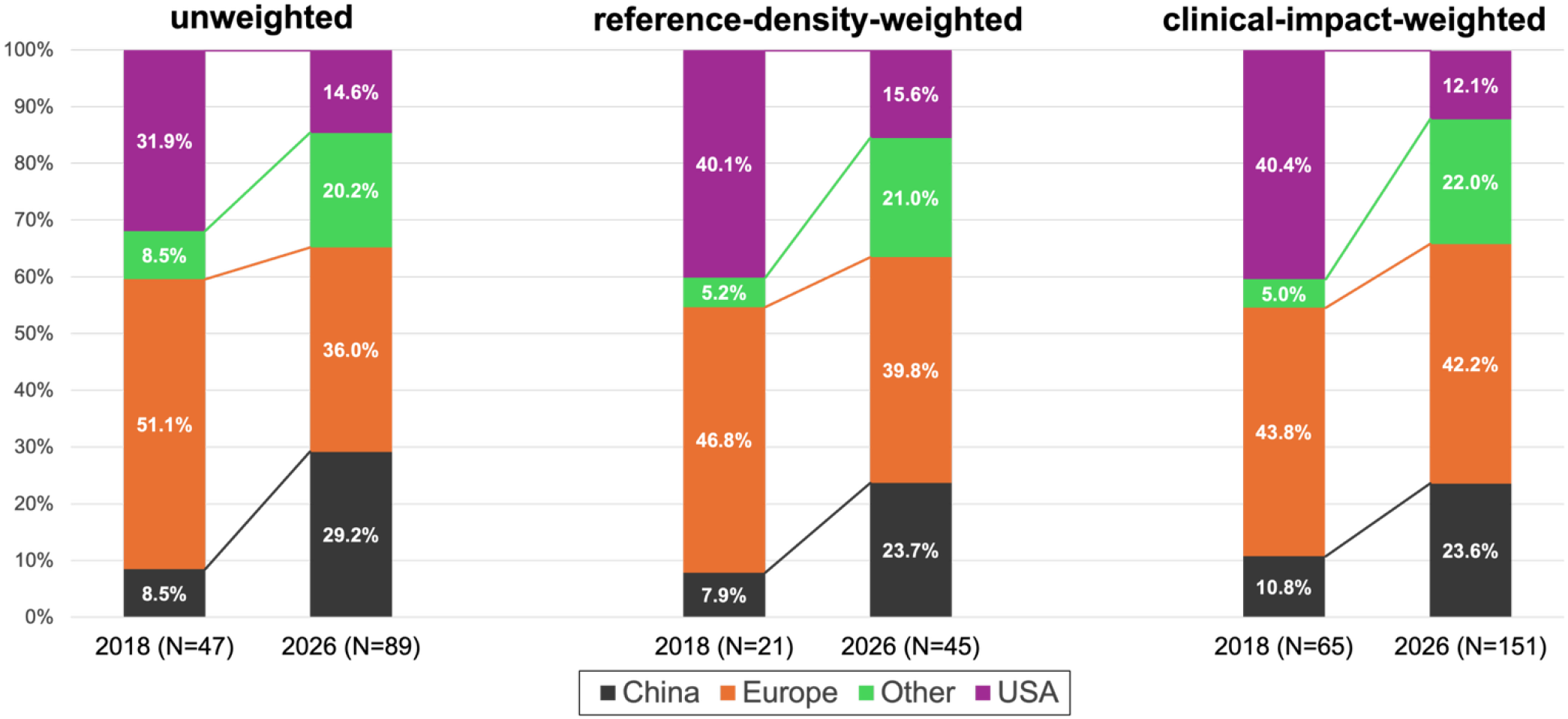
Geographic distribution of randomized controlled trials (RCTs) cited in the 2018 and 2026 American Heart Association/American Stroke Association (AHA/ASA) Acute Ischemic Stroke (AIS) guidelines. Stacked bar graphs show the distribution of RCTs cited in the 2018 and 2026 guidelines across three analytic approaches: unweighted analysis, reference-density-weighted analysis, and clinical-impact-weighted analysis. Within each analytic approach, bars represent the percentage contribution of trials from the China, Europe, United States, and other regions. “Other” includes trials conducted in countries outside the United States, China, and Europe (Australia, New Zealand, Japan, South Korea, Canada, Chile). Differences between guideline eras were statistically significant (unweighted analysis: p=0.002, reference-density-weighted analysis: p=0.003, and clinical-impact-weighted analysis: p=0.002). Percentages were calculated using the number of RCT references in each category (numerator) divided by the total number of RCT references supporting new recommendations for that year and analytic approach (denominator): unweighted (2018: N = 47; 2026: N = 89), reference-density-weighted (2018: N = 21; 2026: N = 45), and clinical-impact-weighted (2018: N = 65; 2026: N = 151).

Research focus shifted significantly over time in unweighted analyses (p<0.01), while weighted distributions were not significant (p>0.05; Figure 2). In 2018, poststroke care was the most common research area (38.3%), followed by imaging (19.2%), EVT and thrombolysis (17% each), and prehospital and systems of care (8.5%). By 2026, EVT (30.3%) and thrombolysis (28.1%) became the top two areas, followed by prehospital and systems of care (19.1%), with poststroke care and imaging dropping to lowest ranks (15.7%, 6.8% respectively). Regional leadership within these research areas also changed markedly (Table 1). The United States decreased proportionally across all research areas, with the most prominent decline in imaging, dropping from the leading contributor (8.5%) in 2018 to 0% in 2026. China increased across almost all research areas except poststroke care, emerging as the top contributor in thrombolysis (0% to 11.2%) and prehospital and systems of care (0% to 6.7%), while Europe retained EVT leadership (8.5% to 12.4%; +3.9%).

**Figure 2.**
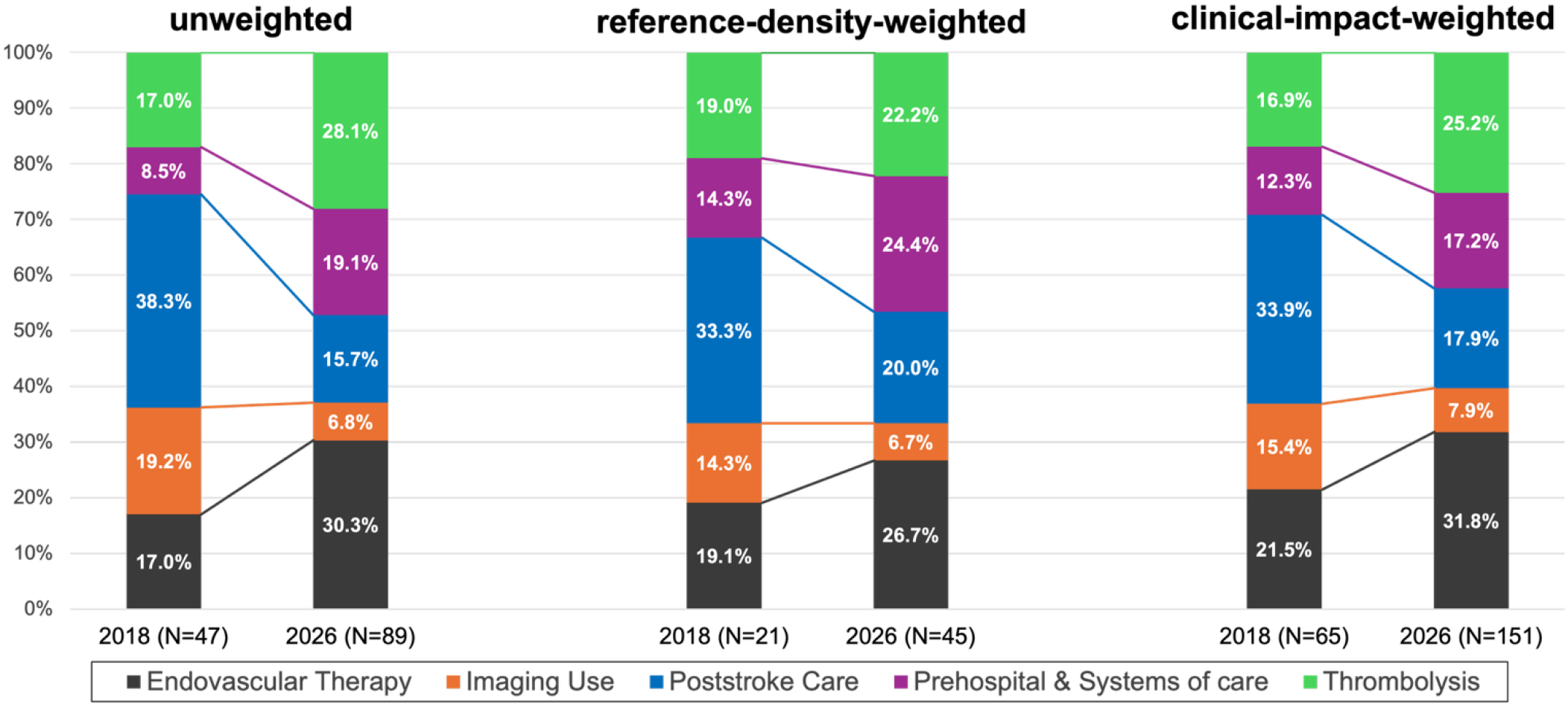
Distribution of RCTs by research area cited in the 2018 and 2026 AHA/ASA AIS guidelines. Stacked bar graphs show the distribution of RCTs cited in the 2018 and 2026 guidelines across three analytic approaches: unweighted analysis, reference-density-weighted analysis, and clinical-impact-weighted analysis. Within each analytic approach, bars represent the percentage of trials in each research area: endovascular therapy, thrombolysis, imaging, poststroke care, and prehospital and systems of care. Differences between guideline eras were significant for the unweighted analysis but not for the weighted analyses (unweighted: p=0.002; reference-density-weighted: p=0.326; clinical-impact-weighted: p=0.274). Percentages were calculated using the number of RCT references in each category (numerator) divided by the total number of RCT references supporting new recommendations for that year and analytic approach (denominator): unweighted (2018: N = 47; 2026: N = 89), reference-density-weighted (2018: N = 21; 2026: N = 45), and clinical-impact-weighted (2018: N = 65; 2026: N = 151).

**Table 1.**
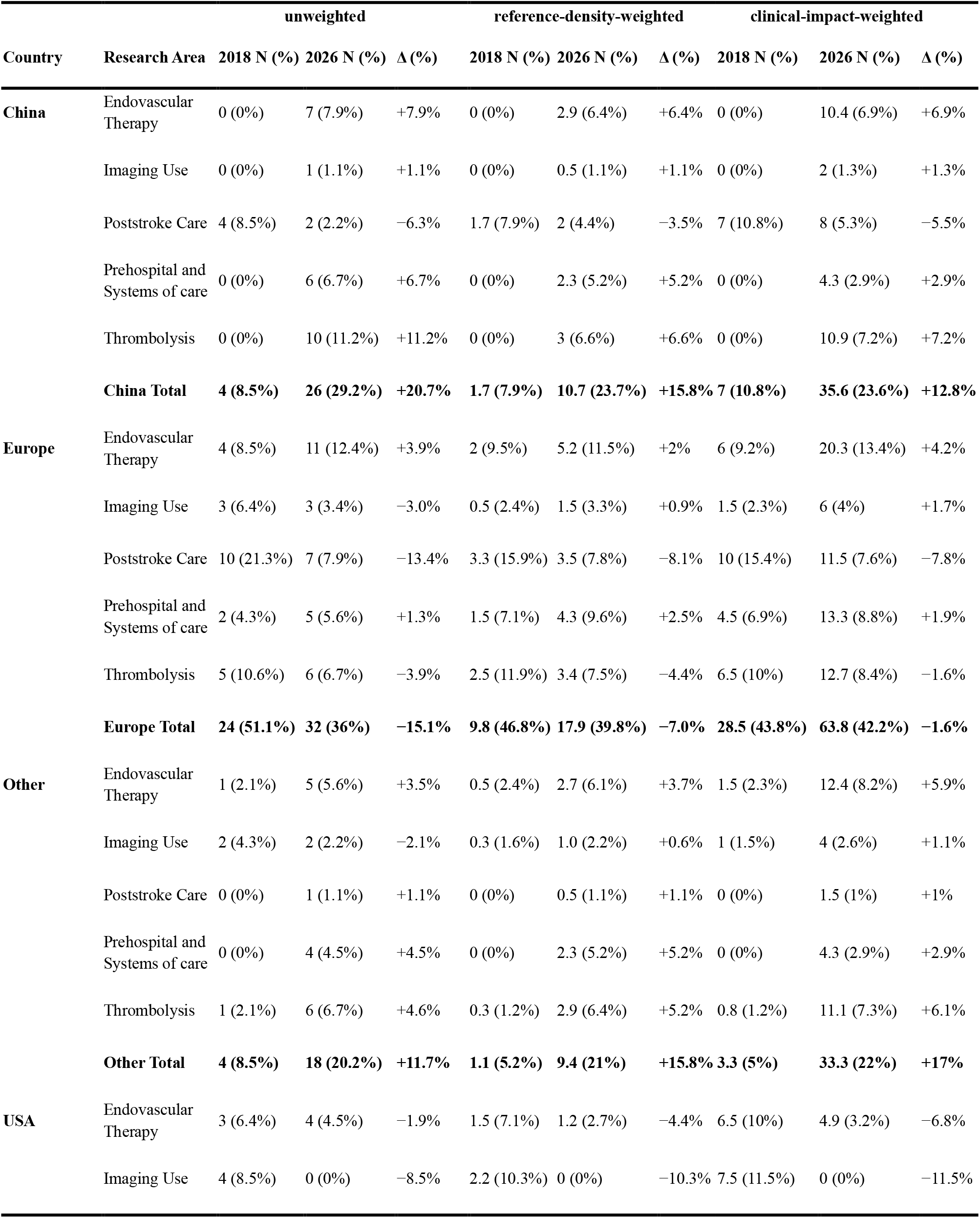
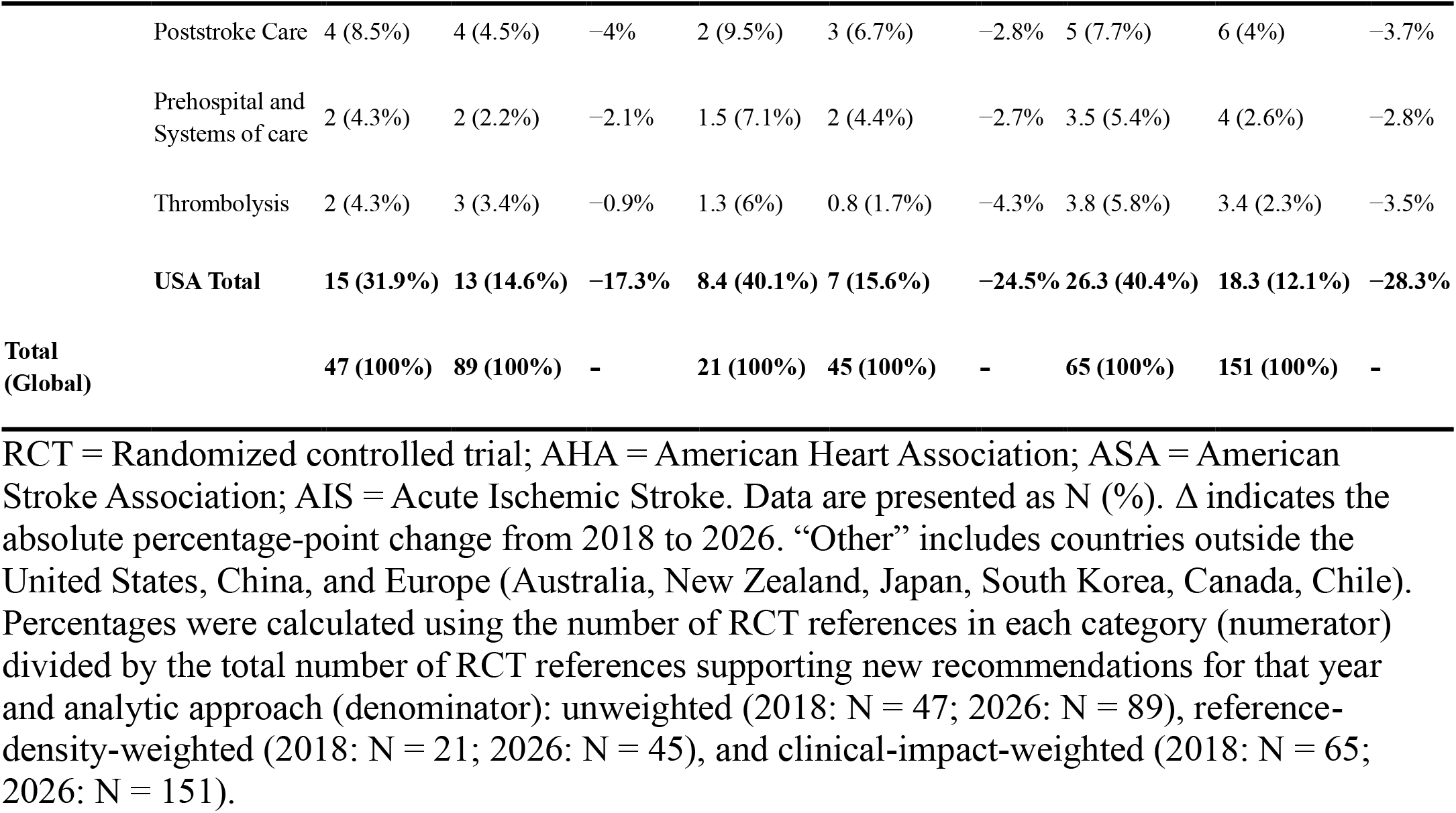
Geographic Distribution of RCTs Supporting New Recommendations in the 2018 and 2026 AHA/ASA AIS Guidelines, Stratified by Research Area.

| Country | Research Area | unweighted |  |  | reference-density-weighted |  |  | clinical-impact-weighted |  |  |
| --- | --- | --- | --- | --- | --- | --- | --- | --- | --- | --- |
|  |  | 2018 N (%) | 2026 N (%) | Δ (%) | 2018 N (%) | 2026 N (%) | Δ (%) | 2018 N (%) | 2026 N (%) | Δ (%) |
| China | Endovascular Therapy | 0 (0%) | 7 (7.9%) | +7.9% | 0 (0%) | 2.9 (6.4%) | +6.4% | 0 (0%) | 10.4 (6.9%) | +6.9% |
|  | Imaging Use | 0 (0%) | 1 (1.1%) | +1.1% | 0 (0%) | 0.5 (1.1%) | +1.1% | 0 (0%) | 2 (1.3%) | +1.3% |
|  | Poststroke Care | 4 (8.5%) | 2 (2.2%) | −6.3% | 1.7 (7.9%) | 2 (4.4%) | −3.5% | 7 (10.8%) | 8 (5.3%) | −5.5% |
|  | Prehospital and Systems of care | 0 (0%) | 6 (6.7%) | +6.7% | 0 (0%) | 2.3 (5.2%) | +5.2% | 0 (0%) | 4.3 (2.9%) | +2.9% |
|  | Thrombolysis | 0 (0%) | 10 (11.2%) | +11.2% | 0 (0%) | 3 (6.6%) | +6.6% | 0 (0%) | 10.9 (7.2%) | +7.2% |
|  | **China Total** | **4 (8.5%)** | **26 (29.2%)** | **+20.7%** | **1.7 (7.9%)** | **10.7 (23.7%)** | **+15.8%** | **7 (10.8%)** | **35.6 (23.6%)** | **+12.8%** |
| Europe | Endovascular Therapy | 4 (8.5%) | 11 (12.4%) | +3.9% | 2 (9.5%) | 5.2 (11.5%) | +2% | 6 (9.2%) | 20.3 (13.4%) | +4.2% |
|  | Imaging Use | 3 (6.4%) | 3 (3.4%) | −3.0% | 0.5 (2.4%) | 1.5 (3.3%) | +0.9% | 1.5 (2.3%) | 6 (4%) | +1.7% |
|  | Poststroke Care | 10 (21.3%) | 7 (7.9%) | −13.4% | 3.3 (15.9%) | 3.5 (7.8%) | −8.1% | 10 (15.4%) | 11.5 (7.6%) | −7.8% |
|  | Prehospital and Systems of care | 2 (4.3%) | 5 (5.6%) | +1.3% | 1.5 (7.1%) | 4.3 (9.6%) | +2.5% | 4.5 (6.9%) | 13.3 (8.8%) | +1.9% |
|  | Thrombolysis | 5 (10.6%) | 6 (6.7%) | −3.9% | 2.5 (11.9%) | 3.4 (7.5%) | −4.4% | 6.5 (10%) | 12.7 (8.4%) | −1.6% |
|  | **Europe Total** | **24 (51.1%)** | **32 (36%)** | **−15.1%** | **9.8 (46.8%)** | **17.9 (39.8%)** | **−7.0%** | **28.5 (43.8%)** | **63.8 (42.2%)** | **−1.6%** |
| Other | Endovascular Therapy | 1 (2.1%) | 5 (5.6%) | +3.5% | 0.5 (2.4%) | 2.7 (6.1%) | +3.7% | 1.5 (2.3%) | 12.4 (8.2%) | +5.9% |
|  | Imaging Use | 2 (4.3%) | 2 (2.2%) | −2.1% | 0.3 (1.6%) | 1.0 (2.2%) | +0.6% | 1 (1.5%) | 4 (2.6%) | +1.1% |
|  | Poststroke Care | 0 (0%) | 1 (1.1%) | +1.1% | 0 (0%) | 0.5 (1.1%) | +1.1% | 0 (0%) | 1.5 (1%) | +1% |
|  | Prehospital and Systems of care | 0 (0%) | 4 (4.5%) | +4.5% | 0 (0%) | 2.3 (5.2%) | +5.2% | 0 (0%) | 4.3 (2.9%) | +2.9% |
|  | Thrombolysis | 1 (2.1%) | 6 (6.7%) | +4.6% | 0.3 (1.2%) | 2.9 (6.4%) | +5.2% | 0.8 (1.2%) | 11.1 (7.3%) | +6.1% |
|  | **Other Total** | **4 (8.5%)** | **18 (20.2%)** | **+11.7%** | **1.1 (5.2%)** | **9.4 (21%)** | **+15.8%** | **3.3 (5%)** | **33.3 (22%)** | **+17%** |
| USA | Endovascular Therapy | 3 (6.4%) | 4 (4.5%) | −1.9% | 1.5 (7.1%) | 1.2 (2.7%) | −4.4% | 6.5 (10%) | 4.9 (3.2%) | −6.8% |
|  | Imaging Use | 4 (8.5%) | 0 (0%) | −8.5% | 2.2 (10.3%) | 0 (0%) | −10.3% | 7.5 (11.5%) | 0 (0%) | −11.5% |

|  |  |  |  |  |  |  |  |  |  |
| --- | --- | --- | --- | --- | --- | --- | --- | --- | --- |
| Poststroke Care | 4 (8.5%) | 4 (4.5%) | −4% | 2 (9.5%) | 3 (6.7%) | −2.8% | 5 (7.7%) | 6 (4%) | −3.7% |
| Prehospital and Systems of care | 2 (4.3%) | 2 (2.2%) | −2.1% | 1.5 (7.1%) | 2 (4.4%) | −2.7% | 3.5 (5.4%) | 4 (2.6%) | −2.8% |
| Thrombolysis | 2 (4.3%) | 3 (3.4%) | −0.9% | 1.3 (6%) | 0.8 (1.7%) | −4.3% | 3.8 (5.8%) | 3.4 (2.3%) | −3.5% |
| **USA Total** | **15 (31.9%)** | **13 (14.6%)** | **−17.3%** | **8.4 (40.1%)** | **7 (15.6%)** | **−24.5%** | **26.3 (40.4%)** | **18.3 (12.1%)** | **−28.3%** |
| **Total (Global)** | **47 (100%)** | **89 (100%)** | **-** | **21 (100%)** | **45 (100%)** | **-** | **65 (100%)** | **151 (100%)** | **-** |

RCT = Randomized controlled trial; AHA = American Heart Association; ASA = American Stroke Association; AIS = Acute Ischemic Stroke. Data are presented as N (%). Δ indicates the absolute percentage-point change from 2018 to 2026. “Other” includes countries outside the United States, China, and Europe (Australia, New Zealand, Japan, South Korea, Canada, Chile). Percentages were calculated using the number of RCT references in each category (numerator) divided by the total number of RCT references supporting new recommendations for that year and analytic approach (denominator): unweighted (2018: N = 47; 2026: N = 89), reference-density-weighted (2018: N = 21; 2026: N = 45), and clinical-impact-weighted (2018: N = 65; 2026: N = 151).

Funding patterns changed more subtly. Overall, government/academic/non-profit funding remained the dominant source in both years (74.5% in 2018 vs. 68.5% in 2026; −6.0%), with a corresponding increase in industry or mixed private funding (25.5% to 31.5%; +6.0%), changes that were not statistically significant (p>0.05 across all weighting schemes; Supplementary Figure 1). When broken down by research areas, government funding grew markedly in prehospital and systems of care (+10.6%) and EVT (+8.4%) but declined sharply in poststroke care (-23.9%), while industry showed gains in thrombolysis (+6.9%) and remained absent from prehospital and systems of care (0% in both guideline years; Table 2). The overall proportion of NIH-funded RCTs was substantial in 2018 (21.3% unweighted, 27.4% reference-density-weighted, 27.3% clinical-impact-weighted), but fell considerably and significantly by 2026 (4.5%, 4.8%, 3.4%, respectively; p<0.01 across all comparisons) (Figure 3).

**Table 2.** Funding Source of RCTs Supporting New Recommendations in the 2018 and 2026 AHA/ASA AIS Guidelines, Stratified by Research Area.

| Funding Source | Research Area | unweighted |  |  | reference-density-weighted |  |  | clinical-impact-weighted |  |  |
| --- | --- | --- | --- | --- | --- | --- | --- | --- | --- | --- |
|  |  | 2018 N (%) | 2026 N (%) | Δ (%) | 2018 N (%) | 2026 N (%) | Δ (%) | 2018 N (%) | 2026 N (%) | Δ (%) |
| **Government / Academic / Non-profit** | Endovascular Therapy | 4 (8.5%) | 15 (16.9%) | +8.4 | 2.2 (10.3%) | 7.1 (15.7%) | +5.4% | 7.8 (12.1%) | 28.8 (19.1%) | +7% |
|  | Imaging Use | 5 (10.6%) | 5 (5.6%) | -5% | 2 (9.5%) | 2.5 (5.6%) | -3.9% | 6.5 (10%) | 10 (6.6%) | -3.4% |
|  | Poststroke Care | 16 (34%) | 9 (10.1%) | -23.9% | 5.8 (27.8%) | 5.5 (12.2%) | -15.6% | 19.5 (30%) | 18.5 (12.3%) | -17.7% |
|  | Prehospital and Systems of care | 4 (8.5%) | 17 (19.1%) | +10.6% | 3 (14.3%) | 11.0 (24.4%) | +10.1% | 8 (12.3%) | 26 (17.2%) | +4.9% |
|  | Thrombolysis | 6 (12.8%) | 15 (16.9%) | +4.1% | 3 (14.3%) | 5.8 (12.8%) | -1.5% | 7 (10.8%) | 23.8 (15.8%) | +5.0% |
|  | **Total** | **35 (74.5%)** | **61 (68.5%)** | **-6%** | **16 (76.2%)** | **31.8 (70.7%)** | **-5.5%** | **48.8 (75.1%)** | **107.1 (70.9%)** | **-4.2%** |
| **Industry (Private / Mixed)** | Endovascular Therapy | 4 (8.5%) | 12 (13.5%) | +5.0% | 1.8 (8.7%) | 4.9 (10.9%) | +2.2% | 6.2 (9.5%) | 19.2 (12.7%) | +3.2% |
|  | Imaging Use | 4 (8.5%) | 1 (1.1%) | -7.4% | 1 (4.8%) | 0.5 (1.1%) | -3.7% | 3.5 (5.4%) | 2 (1.3%) | -4.1% |
|  | Poststroke Care | 2 (4.3%) | 5 (5.6%) | +1.3% | 1.2 (5.6%) | 3.5 (7.8%) | +2.2% | 2.5 (3.8%) | 8.5 (5.6%) | +1.8% |
|  | Prehospital and Systems of care | 0 (0%) | 0 (0%) | 0% | 0 (0%) | 0 (0%) | 0% | 0 (0%) | 0 (0%) | 0% |
|  | Thrombolysis | 2 (4.3%) | 10 (11.2%) | +6.9% | 1 (4.8%) | 4.2 (9.4%) | +4.6% | 4 (6.2%) | 14.2 (9.4%) | +3.2% |
|  | **Total** | **12 (25.5%)** | **28 (31.5%)** | **+6%** | **5 (23.8%)** | **13.2 (29.3%)** | **+5.5%** | **16.2 (24.9%)** | **43.9 (29.1%)** | **+4.2%** |
| **Total (Global)** |  | **47 (100%)** | **89 (100%)** | **-** | **21 (100%)** | **45 (100%)** | **-** | **65 (100%)** | **151 (100%)** | **-** |

Data are presented as N (%). Δ indicates the absolute percentage-point change from 2018 to 2026. Percentages were calculated using the number of RCT references in each category (numerator) divided by the total number of RCT references supporting new recommendations for that year and analytic approach (denominator): unweighted (2018: N = 47; 2026: N = 89), reference-density-weighted (2018: N = 21; 2026: N = 45), and clinical-impact-weighted (2018: N = 65; 2026: N = 151).

**Figure 3.**
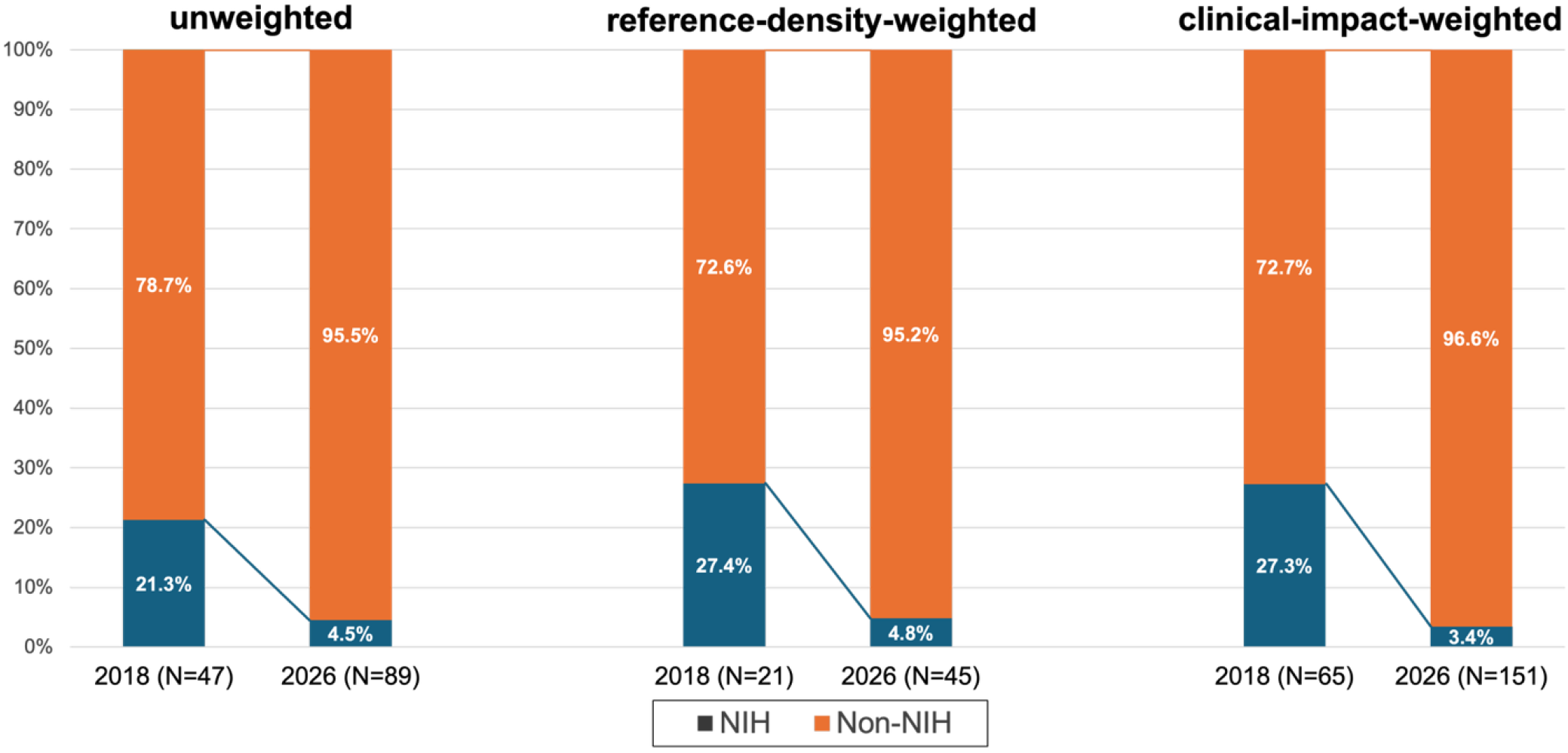
Distribution of RCT funding by NIH vs non-NIH sources cited in the 2018 and 2026 AHA/ASA AIS guidelines. Stacked bar graphs show the proportion of RCTs funded by NIH versus non-NIH sources globally for the 2018 and 2026 guidelines across three analytic approaches: unweighted analysis, reference-density-weighted analysis, and clinical-impact-weighted analysis. Overall, NIH-funding distributions differed significantly between guideline eras (unweighted: p=0.002; reference-density-weighted: p=0.002; clinical-impact-weighted: p<0.001). Percentages were calculated using the number of RCT references in each category (numerator) divided by the total number of RCT references supporting new recommendations for that year and analytic approach (denominator): unweighted (2018: N = 47; 2026: N = 89), reference-density-weighted (2018: N = 21; 2026: N = 45), clinical-impact-weighted (2018: N = 65; 2026: N = 151).

## DISCUSSION

In this analysis of global trends in practice-changing AIS research using the AHA/ASA AIS guidelines, we observed a clear shift in geographic distribution and funding patterns. Europe remained the largest source of RCT citations. On the other hand, whereas U.S.-based research had led to the second highest number of citations for new recommendations in the 2018 guidelines, it contributed the least of the 4 regions in 2026, and the U.S. was the only one to have significantly decreasing impact in all 3 weighting approaches. In addition, whereas NIH-funded RCTs played an outsized role on new recommendations in 2018 (nearly one-fourth of all the new recommendations), its impact fell to < 5% in 2026. Conversely, Chinese research became the second most cited source of evidence (nearly one-third of all references), with growth in nearly all the AIS research topics.

For many years, trials from China and other rapidly expanding research regions had been viewed with caution. Several studies have raised concerns about data accuracy, including documented higher retraction rates, incomplete trial registration, selective reporting, language and indexing biases, and systemic pressures to produce positive results.^10–14^ However, similar issues have been reported in other international datasets.^15,16^ In addition, differences in ethnic diversity, stroke etiology (i.e. greater rates of hemorrhagic stroke, intracranial atherosclerosis), and healthcare systems had led to questions around generalizability to U.S.-based populations.^17–20^ These issues had previously influenced how researchers evaluate studies and uphold standards of research integrity and external validity.^21^ However, the current analysis reveals that several of the most high-impact 2026 recommendations now rest primarily on non-U.S. RCTs. A prominent example is basilar artery occlusion, where the Class I recommendation for EVT within 24 hours is built on Chinese multicenter trials such as ATTENTION and BAOCHE.^22,23^ These trials were conducted in large, specialized stroke centers with high patient volumes and centralized systems that facilitate rapid enrollment and execution. In this context, while concerns of Chinese RCT validity and generalizability may have previously led to hesitancy in their use for practice guideline generation, that reluctance may no longer be sustainable given the difficulty of generating comparable U.S.-based evidence for such time-sensitive interventions. It may be more productive to consider these trials within the same methodological and ethical framework applied to trials in other regions, while also recognizing that structural and demographic factors, such as larger at-risk populations and efficient care networks, may facilitate large-scale, high-quality AIS research in certain settings.

The decline in NIH-funded clinically impactful research is particularly notable. In 2018, NIH-supported trials, such as NINDS rt-PA, INSTINCT, and NIH-backed components of DEFUSE 3, accounted for roughly one-quarter of guideline-driving RCTs globally and two-thirds of U.S.-origin trials.^24–26^ By 2026, citations to NIH-funded trials dropped across all weighting strategies, despite the continued operation of StrokeNet, established in 2013 to facilitate multicenter stroke research. On NIH RePORTER, the total funding for StrokeNet has not been substantially reduced from the period of 2018 to 2025, averaging around $30 million per year throughout this time period.^27^

At the same time, structural and systemic factors may further limit the clinical impact of U.S.-based stroke trials. Registry-based analyses indicate that, although the United States continues to register the largest number of clinical trials globally, including in AIS, growth in AIS trials has been more modest compared with other fields and remains unevenly distributed within the country, with several high-burden regions like Mississippi and Louisiana underrepresented.^7,9^ The sharper decline in U.S.-based RCT impact under clinical-impact-weighting may reflect that many U.S./NIH stroke trials may have been duplicated in other settings or that these U.S.-based studies targeted less impactful questions. These structural imbalances, combined with rising trial costs, regulatory complexity, and recruitment challenges, particularly in time-sensitive AIS populations, may further limit the efficiency and clinical impact of U.S.-based AIS trials. Differences in population size and healthcare system structure may also contribute to more rapid patient recruitment and trial completion in some international settings, where centralized systems and larger patient volumes facilitate large-scale enrollment.^28^ These issues may continue to compound, given the substantially lower trial costs outside North America (49–78% of U.S. costs per site).^29–32^

This shift carries immediate implications for U.S. practice. Clinicians now rely heavily on non-U.S. trials for routine decisions about reperfusion therapies, patient selection, and systems of care. While U.S. clinicians benefit from expanded high-quality global evidence, reduced domestic representation within guideline-defining research likely reflects these structural and competitive factors. Reference selection in guidelines is inherently subjective, with authors often favoring studies from their own institutions, collaborators, or home countries.^33,34^ In our dataset, both the 2018 and 2026 AIS guidelines were written by predominantly U.S.-based groups, conditions under which selection bias would most likely favor U.S. and NIH-funded trials.^35^ Yet U.S.-origin RCTs lost ground across all weighted analyses, and the relative influence of NIH-funded trials dropped even more steeply. This finding suggests that the true global shift away from U.S. centrality is likely even larger than what we measure using guideline citations alone. Improving the translational efficiency of NIH- and StrokeNet-supported research will be critical to preserving U.S. leadership in evidence generation, by prioritizing high-impact trials that are both feasible and positioned to meaningfully change care and compete globally.^36^ This shift also underscores that no single country should be assumed to be the default reference population, and that guideline-informing evidence is increasingly drawn from diverse global settings.

There are limitations to this study that should be acknowledged. First, we analyzed only guideline-cited RCTs, even though high-quality observational cohorts, registry data, and mechanistic studies also influence recommendations. We made this choice intentionally. Because non-RCTs derive lower classes of recommendations, and the purpose of our study was to study practice-changing research, these other types of study designs were excluded. Next, we examined only two guideline time points, which provides a snapshot rather than continuous trajectory across the intervening years. However, these two guidelines represent RCTs that span over a decade and likely capture longer-term trends. Third, we did not formally assess trial quality or risk of bias, relying instead on guideline inclusion as our quality filter. We also did not evaluate research integrity issues by country or sponsor. Finally, our broad funding categories (NIH vs non-NIH; government/academic/non-profit vs industry) simplify complex funding arrangements and may mask important differences in how public and private entities collaborate on individual trials. Nevertheless, RCTs in AHA guidelines are not random; they represent the studies that expert groups judge to be most relevant for defining standard of care and often influence certification requirements, quality metrics, and reimbursement decisions.

In this bibliometric analysis of the AHA/ASA AIS guidelines, we identified a large contribution of clinical-practice changing U.S.-based research and NIH-funded RCTs in 2018, which fell considerably in 2026. We also identified a converse increase in RCTs from China. Europe remained the dominant region. These findings highlight the need to reconsider our approach to AIS clinical research in the U.S. and to counteract the declining clinical impact of research outputs.

## Data Availability

All data used in this study are derived from publicly available sources and are included with the manuscript and supplementary materials.

## Acknowledgments

None

